# A Bayesian Bivariate Spatial Analysis of the Shared and Distinct Determinants of Stunting and Wasting Among Children in Ethiopia: Evidence from the 2019 Mini DHS

**DOI:** 10.64898/2026.02.19.26346605

**Authors:** Yilikal Tesfaye Haile

## Abstract

Childhood malnutrition remains a major public health challenge in Ethiopia, where stunting and wasting co-exist but may arise from distinct spatial and etiological processes. Analyses focusing on a single outcome may overlook the interdependence of these conditions and their geographic heterogeneity. This study aimed to disentangle the determinants of stunting and wasting among children under five years of age using a Bayesian bivariate spatial modelling framework. Data from 5,405 children included in the 2019 Ethiopia Mini Demographic and Health Survey were analyzed. Stunting and wasting were modelled as correlated binary outcomes using Bayesian bivariate hierarchical geostatistical models implemented through SPDE-INLA, accounting for child, maternal, household, and environmental covariates, non-linear age effects, and spatial dependence. Model performance was assessed using the deviance information criterion, Watanabe–Akaike information criterion, and marginal log-likelihood. The bivariate model identified shared socio-economic and biological determinants. Multiple births, male sex, low maternal education, a higher number of under-five children, and household poverty were associated with increased risks of both outcomes. Female-headed households were associated with lower odds of stunting but higher odds of wasting. Spatial analysis revealed elevated residual stunting risk in the northern and central highlands, whereas wasting hotspots were concentrated in northeastern pastoralist regions. Residual spatial correlation was weak (ρ = −0.12), indicating largely independent geographic patterns. These findings suggest that effective child nutrition policies in Ethiopia require outcome-specific and regionally tailored interventions addressing both chronic and acute forms of malnutrition.

## Introduction

Childhood malnutrition is not merely a health statistic; it is a profound barrier to global human capital. While the world struggles with 150.2 million stunted and 42.8 million wasted children, the crisis has found a relentless epicenter in Africa. As of 2024, approximately 64.8 million children under five on the continent are affected by stunting, accounting for nearly 43% of the global total. Critically, Africa remains the only region where the absolute number of stunted children continues to rise (1).

Within this landscape, Ethiopia stands at a critical juncture. Despite sustained policy efforts, the nation faces a dual burden: a staggering 37–42% stunting rate and acute wasting affecting up to 11% of its children under five (2). These figures represent a collision of chronic structural deprivation and recurrent environmental shocks. To secure Ethiopia’s future, we must move beyond generalized solutions and pivot towards context-specific, evidence-based interventions that address these interconnected pathways of malnutrition.

Recent evidence suggests that stunting (chronic) and wasting (acute) are far from independent conditions; they are biologically and behaviorally intertwined (3). Biologically, stunting often represents a deleterious physiological adaptation to repeated episodes of wasting. When a child experiences acute weight loss, the body’s metabolic response halts linear growth to conserve energy for vital survival functions (3). Behaviorally, these outcomes are linked through shared risk factors such as poor infant and young child feeding practices and recurrent infections like diarrhea, which drive a vicious cycle where the wasting of today becomes the stunting of tomorrow (4).

Despite this biological reality, most existing studies in Ethiopia treat stunting and wasting as independent, siloed outcomes, which fails to account for their intrinsic correlation and leads to a fragmented understanding of a child’s true status (5–7). Furthermore, non-spatial models often ignore spatial autocorrelation the neighborhood effect whereby nearby areas share similar risks due to climate, food markets, or health infrastructure (8–10). Ignoring these dependencies can lead to biased parameter estimates and mask critical hidden hotspots at the sub regional level (11).

This study addresses these gaps by adopting a bivariate spatial modelling framework. First, modelling outcomes simultaneously allows the framework to borrow strength, leveraging information from the more stable indicator to improve the precision of estimates for the more volatile one (12). Second, it allows for the identification of shared risk surfaces by quantifying a shared spatial component, revealing unobserved environmental factors that fuel both conditions (13). Finally, a bivariate approach estimates the residual correlation between stunting and wasting, allowing us to determine where these conditions are coupled and where they are decoupled due to localized agro-ecological or socio-economic shocks (14).

Utilizing Bayesian Bivariate Spatial (SPDE-INLA) models, this study provides a high-resolution diagnostic tool to decouple the drivers of malnutrition across Ethiopia’s diverse landscape, moving beyond national averages towards targeted, evidence-based policy.

## Materials and Methods

### Study Design

This study employed a cross-sectional, population-based design using secondary data to examine the spatial distribution and determinants of stunting and wasting among children under five years of age in Ethiopia. The analysis was conducted at the individual child level, with children nested within households and geographic clusters. While the underlying survey design assumes independent sampling units, spatial dependence and outcome correlation were explicitly addressed during the statistical analysis through a Bayesian hierarchical geostatistical modelling framework (15,16).

### Data Source and Study Population

The data for this study were secondary and extracted from the 2019 Ethiopia Mini Demographic and Health Survey (EMDHS), accessed on 17 November 2025 after obtaining permission from the The DHS Program. The EMDHS is a nationally representative survey implemented by the Ethiopian Public Health Institute in collaboration with the Central Statistical Agency(17). The survey utilized a two-stage stratified cluster sampling design (18). In the first stage, 305 Enumeration Areas (EAs) (93 in urban and 212 in rural areas) were selected with probability proportional to EA size from the 2019 national census frame. In the second stage, a systematic sample of households was selected from each EA. The study population consisted of all children aged 0–59 months residing in the sampled households who had valid anthropometric measurements. After cleaning for missing values and outliers, the final analytical sample included 5,405 children from 305 clusters. The author did not have access to any information that could identify individual participants either during or after data collection, as the dataset was fully anonymized prior to release.

### Study Variables

The study variables were categorized into bivariate outcomes and a set of multi-level explanatory predictors. The dependent variables were stunting and wasting, treated as binary responses. Following the World Health Organization (WHO) Child Growth Standards, stunting was defined as a Height-for-Age Z-score (HAZ) < -2.0 standard deviations (SD) from the median of the reference population, representing chronic growth failure. Wasting was defined as a Weight-for-Height Z-score (WHZ) < -2.0 standard deviations SD, representing acute nutritional stress (19).

Predictors were selected based on their established relevance in the UNICEF framework for malnutrition(20). Child-level variables included age in months, sex, breastfeeding status (breastfed, never breastfed), Birth spacing (first birth, < 24 months, and > 24 months), and plurality of birth (single vs. multiple). Household and maternal variables included number of children aged under five years (one, two, and above two), sex of household head, household wealth index (poorest, poorer, middle, richer, richest), maternal education (no education, primary, secondary, or higher), place of delivery (home vs health institution) and maternal age. Environmental and structural variables included the place of residence (urban vs. rural) and geographic location (longitude and latitude of the cluster).

### Statistical Analysis and Model Specification

A Bayesian bivariate hierarchical geostatistical model was used to jointly analyze stunting and wasting as correlated binary outcomes. Joint modelling allows simultaneous estimation of shared and outcome-specific covariate effects while accounting for residual correlation between chronic and acute forms of undernutrition, thereby improving efficiency and interpretability compared with separate univariate models (21). This framework allow us to estimate the residual spatial correlation (*ρ*), which quantifies the degree to which unexplained risks for both conditions overlap geographically

Let 𝑦_𝑖𝑗_^(𝑘)^ ∈ {0,1} denote the nutritional outcome k (k= 1, stunting and k = 2, wasting) for child ***i*** residing in cluster ***j***. Conditional on the latent effects, outcomes were assumed to follow a Bernoulli distribution:

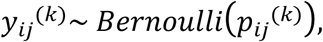

With logit link function

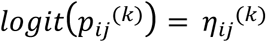

The linear predictor was specified as

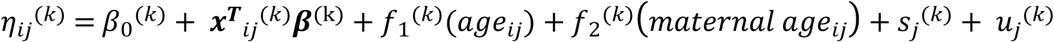

Where 𝛽_0_^(𝑘)^is an outcome-specific intercepts and 𝒙_𝒊𝒋_denotes the vector of observed categorical predictors. All covariates were modelled with outcome-specific effects, allowing their associations to differ between chronic and acute undernutrition. Continuous covariates child age and maternal age at birth were modelled using second-order random walk (RW2) priors to flexibly capture non-linear relationships consistent with childhood growth-faltering patterns (22). Penalized complexity (PC) priors were assigned to precision parameters to control model complexity. To account for residual correlation among children sampled within the same enumeration area, cluster-level random effects were included as independent and identically distributed Gaussian terms. These were specified with an exchangeable correlation structure across outcomes, permitting stunting and wasting to share unobserved cluster-level influences while retaining outcome-specific variability (23).

Spatial dependence was captured through outcome-specific spatial random fields 𝑠_𝑗_^(𝑘)^, defined over the geographic coordinates of DHS clusters. The spatial effects were modelled using the stochastic partial differential equation (SPDE) approach, which represents a continuously indexed Gaussian random field with a Matérn covariance function as a Gaussian Markov random field over a triangulated mesh (24).

The Matérn covariance function is given by

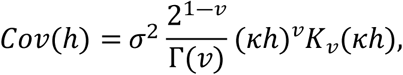

Where h is the distance between locations, 𝜎^2^ is the marginal variance, *ν* controls smoothness, and 𝜅 is related to the spatial range. The SPDE formulation enables efficient computation and inference for large spatial datasets. Correlation between the spatial fields of stunting and wasting was modelled using an exchangeable multivariate structure, allowing identification of shared geographic risk patterns (13,24).

To account for the complex survey design of the Ethiopia Demographic and Health Survey (EDHS), including unequal selection probabilities and non-response, sampling weights were incorporated at the individual child level. Weights were normalized to preserve the effective sample size and entered through a weighted likelihood formulation within the Bayesian framework (25,26).

Bayesian inference was performed using Integrated Nested Laplace Approximation (INLA), which provides accurate and computationally efficient approximations to posterior marginal for latent Gaussian models (27). Weakly informative penalized complexity (PC) priors were assigned to precision parameters of random effects to control model complexity and prevent overfitting(28). Model adequacy was assessed using the deviance information criterion (DIC), Watanabe–Akaike information criterion (WAIC), and Marginal log-Likelihood.

All statistical analyses were conducted in R software version 4.5.0. Bayesian hierarchical models were fitted using the Integrated Nested Laplace Approximation (INLA) approach, implemented with the R-INLA package (version 25.11.12.1). Spatial random fields were constructed using a triangulated mesh for the SPDE approach, and all post-processing, including extraction of posterior summaries and visualization of spatial risk surfaces, was performed within R.

## RESULTS

### Descriptive and Bivariate Analysis

The study population consisted of 5,405 children under the age of five with complete anthropometric and covariate data. The descriptive characteristics of the sample, stratified by stunting and wasting status, are detailed in Table 1. Pearson’s chi-square tests were employed to assess bivariate associations between the outcomes and the hypothesized predictors.

**Table 1:** Bivariate association between child, maternal, and household-level characteristics and the malnutrition status (stunting and wasting) of children under five in Ethiopia (n = 5,405)

| Variable | Category | Stunted |  |  | Wasted |  |  |
| --- | --- | --- | --- | --- | --- | --- | --- |
|  |  | yes | no | p-value | Yes | No | p-value |
| Child Sex | Male | 976 (35.2%) | 1797 (64.8%) | 0.0075 | 283 (10.2%) | 2490 (89.8%) | 0.0002 |
|  | Female | 835 (31.7%) | 1797 (68.3%) |  | 191 (7.3%) | 2441 (92.7%) |  |
| Plurality of birth | Single | 1758 (33.2%) | 3533 (66.8%) | 0.0041 | 459 (8.7%) | 4832 (91.3%) | 0.1320 |
|  | Multiple | 53 (46.5%) | 61 (53.5%) |  | 15 (13.2%) | 99 (86.8%) |  |
| Breastfed | breastfed | 1718 (33.2%) | 3455 (66.8%) | 0.0358 | 446 (8.6%) | 4727 (91.4%) | 0.0896 |
|  | never breastfed | 93 (40.1%) | 139 (59.9%) |  | 28 (12.1%) | 204 (87.9%) |  |
| Birth spacing | First birth | 344 (29.3%) | 831 (70.7%) | <0.0001 | 88 (7.5%) | 1087 (92.5%) | 0.161 |
|  | <24 months | 377 (38.2%) | 610 (61.8%) |  | 96 (9.7%) | 891 (90.3%) |  |
|  | >=24 months | 1090 (33.6%) | 2153 (66.4%) |  | 290 (8.9%) | 2953 (91.1%) |  |
| Number of under 5 children | one | 618 (30.1%) | 1435 (69.9%) | <0.0001 | 141 (6.9%) | 1912 (93.1%) | 0.0002 |
|  | two | 879 (36.4%) | 1534 (63.6%) |  | 229 (9.5%) | 2184 (90.5%) |  |
|  | three and more | 314 (33.4%) | 625 (66.6%) |  | 104 (11.1%) | 835 (88.9%) |  |
| Place delivery | Health institution | 651 (33.4%) | 1297 (66.6%) | 0.943 | 125 (6.4%) | 1823 (93.6%) | <0.0001 |
|  | Home | 1160 (33.6%) | 2297 (66.4%) |  | 349 (10.1%) | 3108 (89.9%) |  |
| Sex of household head | Male | 1513 (35.1%) | 2798 (64.9%) | <0.0001 | 342 (7.9%) | 3969 (92.1%) | <0.0001 |
|  | Female | 298 (27.2%) | 796 (72.8%) |  | 132 (12.1%) | 962 (87.9%) |  |
| Maternal Education | No education | 1125 (38%) | 1832 (62%) | <0.0001 | 329 (11.1%) | 2628 (88.9%) | <0.0001 |
|  | Primary | 554 (32.7%) | 1142 (67.3%) |  | 112 (6.6%) | 1584 (93.4%) |  |
|  | Secondary | 90 (19.5%) | 372 (80.5%) |  | 22 (4.8%) | 440 (95.2%) |  |
|  | Higher | 42 (14.5%) | 248 (85.5%) |  | 11 (3.8%) | 279 (96.2%) |  |
| Wealth Index | Poorest | 679 (37.1%) | 1151 (62.9%) | <0.0001 | 262 (14.3%) | 1568 (85.7%) | <0.0001 |
|  | Poorer | 358 (38.5%) | 573 (61.5%) |  | 76 (8.2%) | 855 (91.8%) |  |
|  | Middle | 293 (38.6%) | 467 (61.4%) |  | 41 (5.4%) | 719 (94.6%) |  |
|  | Richer | 241 (34.4%) | 460 (65.6%) |  | 41 (5.8%) | 660 (94.2%) |  |
|  | Richest | 240 (20.3%) | 943 (79.7%) |  | 54 (4.6%) | 1129 (95.4%) |  |
| Place of Residence | Urban | 279 (22.3%) | 972 (77.7%) | <0.0001 | 77 (6.2%) | 1174 (93.8%) | 0.0002 |
|  | Rural | 1532 (36.9%) | 2622 (63.1%) |  | 397 (9.6%) | 3757 (90.4%) |  |

The results indicate that child sex is significantly associated with both nutritional outcomes, with a higher prevalence of stunting (35.2%; *P* = 0.0075) and wasting (10.2%; *P* = 0.0002) observed among male children. Biological and feeding factors showed varied impacts; while plurality of birth (*P* = 0.0041) and breastfeeding status (*P* = 0.0358) were significantly associated with chronic growth failure, they did not reach the threshold for statistical significance for acute malnutrition at the 5% level. Similarly, birth spacing was a highly significant predictor for stunting (*P* < 0.0001) but not for wasting (*P* = 0.161).

Socio-economic determinants exhibited a robust and statistically significant association with both outcomes (*P* < 0.0001 for all measures). A clear inverse gradient was observed between maternal education and malnutrition; stunting prevalence declined monotonically from 38.0% among children of mothers with no formal education to 14.5% for those with higher education. A parallel trend was observed for household wealth, where the risk of both stunting and wasting was markedly lower in the richest quintile compared to the poorest.

Significant geographic and household-level disparities were also identified. Children residing in rural areas faced a significantly higher burden of stunting (36.9%) and wasting (9.6%) compared to their urban counterparts (*P <* 0.0002). Furthermore, the sex of the household head was strongly associated with both outcomes (*P* < 0.0001); notably, children in female-headed households presented a lower prevalence of stunting (27.2%) but a higher prevalence of wasting (12.1%) relative to male-headed households. Amusingly, place of delivery showed no significant association with stunting (*P* = 0.943) but was highly significant for wasting (*P* < 0.0001), with home deliveries associated with a higher prevalence of acute malnutrition.

### Exploratory Spatial Analysis of Observed Prevalence

The empirical spatial distribution of observed cluster-level prevalence for stunting and wasting (Figure 1) reveals significant geographic heterogeneity across Ethiopia. For stunting (a), the raw data shows a dense concentration of high-prevalence clusters throughout the northern and central highlands, particularly within the Amhara and Tigray regions, where numerous clusters exceed a 40% prevalence rate. In contrast, the spatial pattern for wasting (b) appears more dispersed, characterised by localized “hotspots” of acute malnutrition. The highest observed wasting prevalence is predominantly clustered in the northeastern pastoralist areas of the Afar region and parts of the Somali region, with several clusters surpassing the 15% critical public health threshold. These observed patterns provide strong initial evidence of spatial non-randomness and clustering, justifying the application of formal Bayesian spatial modelling to account for the underlying spatial dependencies and to derive smoothed, reliable estimates of nutritional risk.

**Figure 1.**
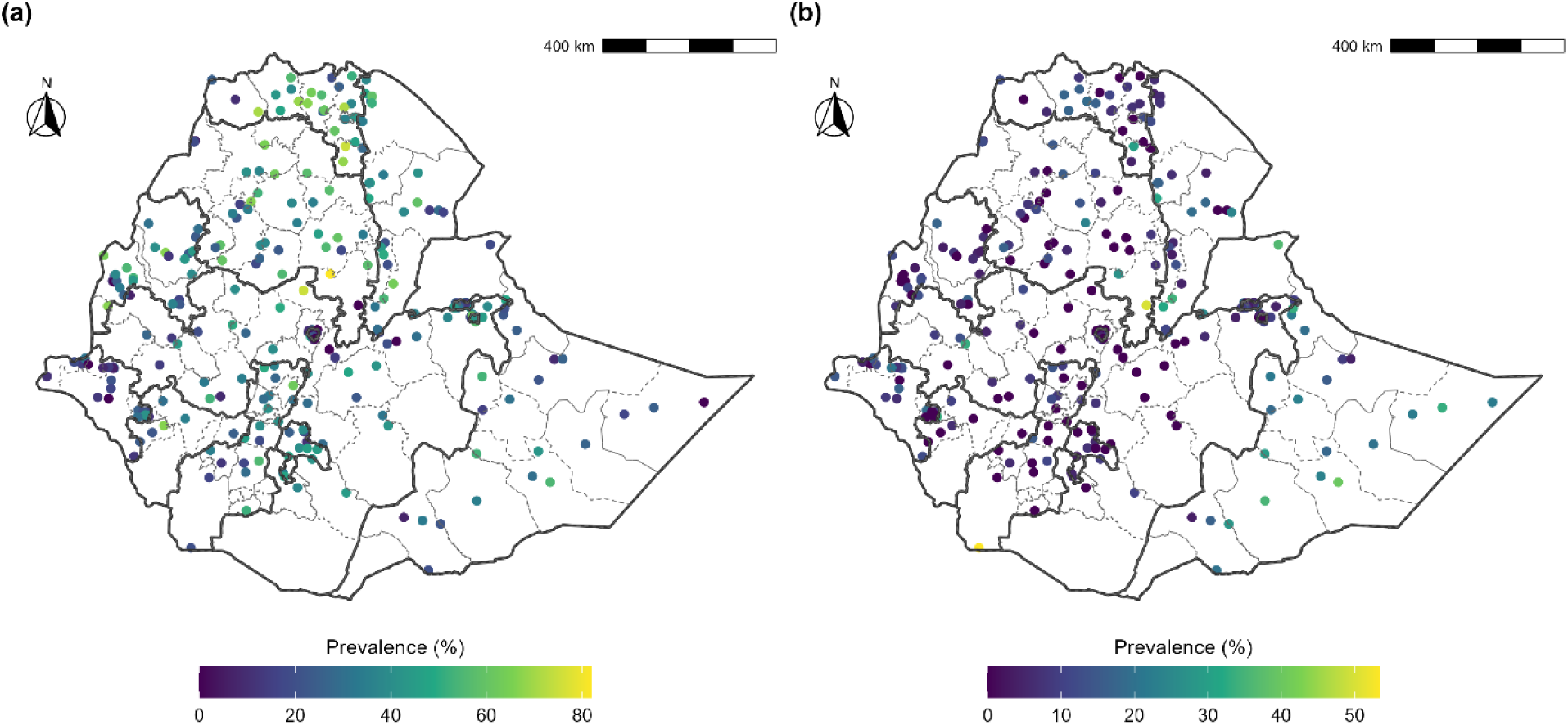
Spatial distribution of observed cluster-level prevalence in Ethiopia a) Stunting and b) Wasting.

### Model Selection and Comparative Performance

To determine the most robust framework for capturing the spatial and structural determinants of childhood malnutrition, four Bayesian hierarchical models were evaluated using the Deviance Information Criterion (DIC), the Watanabe-Akaike Information Criterion (WAIC), and the marginal log-likelihood (Table 2). Model 1, which assumed a shared spatial effect between stunting and wasting, exhibited the highest (least favorable) information criteria (DIC: 9078.57; WAIC: 9298.21). Transitioning to a bivariate spatial framework (Model 2) which allows for outcome-specific spatial random effects resulted in a substantial reduction in DIC (Δ DIC = 159.36), confirming that stunting and wasting follow distinct, although potentially correlated, geographic processes. The inclusion of nonlinear effects further enhanced model performance. The addition of a smooth term for child age (Model 3) significantly improved the model fit, and the final specification (Model 4), which incorporated nonlinear functions for both child age and maternal age at birth alongside the bivariate spatial structure, achieved the lowest DIC (8753.63) and WAIC (9114.06). Furthermore, Model 4 yielded the highest marginal log-likelihood (−4645.46), providing definitive evidence that the integration of bivariate spatial dependencies and nonlinear covariates offers the most parsimonious and accurate representation of the nutritional data. Consequently, all subsequent spatial predictions and posterior inferences are based on the results of Model 4.

**Table 2:** Bayesian model comparison for bivariate spatial models of childhood stunting and wasting in Ethiopia (EDHS 2019)

| Model | Description | DIC | WAIC | Marginal log-Likelihood |
| --- | --- | --- | --- | --- |
| Model 1 | Shared spatial effect | 9078.57 | 9298.21 | -4686.09 |
| Model 2 | Bivariate spatial effect | 8919.21 | 9223.57 | -4711.94 |
| Model 3 | Bivariate spatial + nonlinear child age | 8792.76 | 9128.64 | -4659.83 |
| Model 4 | Bivariate spatial + nonlinear child age and maternal age | 8753.63 | 9114.06 | -4645.46 |

### Spatial Hyper parameters and Process Characteristics

The posterior summaries for the spatial hyper parameters of the bivariate SPDE model are presented in Table 3. The estimated nominal spatial range was 158 km (95% CI: 102 – 233 km). This parameter signifies the distance at which the spatial correlation between locations becomes statistically negligible, suggesting that the unobserved environmental and socio-economic drivers of malnutrition in Ethiopia operate over a relatively broad regional scale rather than being strictly localized to individual villages.

**Table 3:** Posterior summaries of spatial hyper parameters.

| Parameter | Posterior mean | SD | 95% CI |
| --- | --- | --- | --- |
| Spatial range (km) | 158 | 32.8 | 102 – 233 |
| Spatial SD | 0.74 | 0.068 | 0.61 – 0.88 |
| Spatial correlation ($\rho$) | -0.12 | 0.154 | -0.41 – 0.19 |

The spatial standard deviation (SD) was estimated at 0.74 (95% CI: 0.61 – 0.88), reflecting a substantial magnitude of residual spatial heterogeneity that remains after adjusting for all observed covariates. Furthermore, the spatial correlation (ρ) between stunting and wasting was found to be -0.12 (95% CI: -0.41 – 0.19). Although the point estimate suggests a weak negative relationship in the residual spatial effects, the credible interval crosses zero, indicating that while the outcomes are modelled jointly, their unexplained geographic patterns are largely independent after accounting for shared risk factors like wealth and maternal education.

### Risk factors of Stunting and Wasting

The posterior odds ratios (OR), standard deviation and 95% credible intervals (CI) from the final bivariate spatial model are summarized in Table 4. Several child, maternal, and household-level characteristics emerged as significant predictors of malnutrition. Child sex was a robust determinant for both outcomes; female children had significantly lower odds of being stunted (OR: 0.71, 95% CI: 0.62–0.80) and wasted (OR: 0.59, 95% CI: 0.47–0.74) compared to males. Plurality of birth also showed a strong association, with children from multiple births facing nearly double the odds of stunting (OR: 1.82, 95% CI: 1.20–2.76) and more than twice the odds of wasting (OR: 2.36, 95% CI: 1.32–4.24) compared to singletons.

**Table 4:**
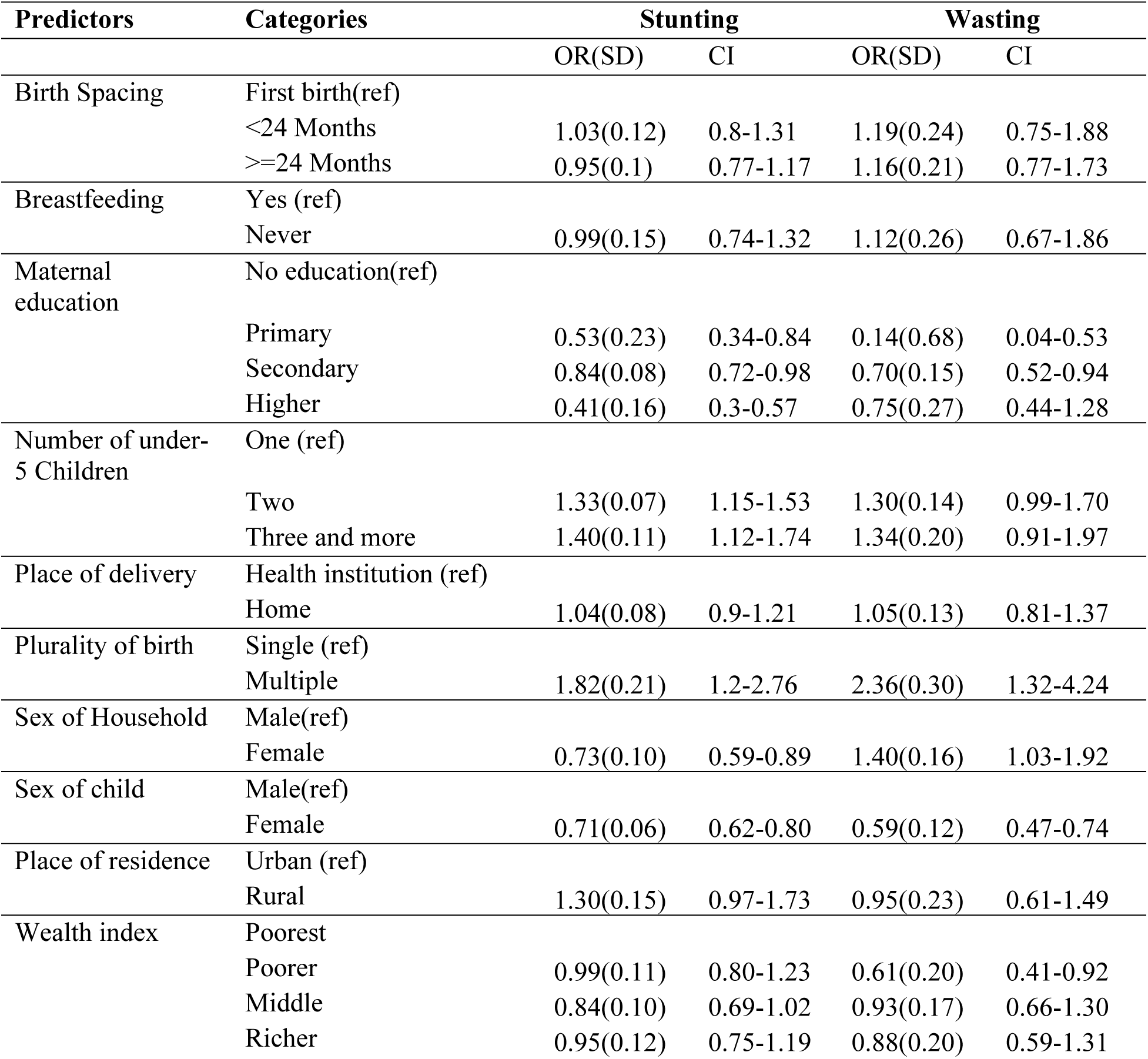
Posterior odds ratios (OR), standard deviations, and 95% credible intervals from the bivariate Bayesian spatial (SPDE) model for stunting and wasting among under-five children, Ethiopia.

Household composition and socio-economic status significantly influenced chronic malnutrition. The odds of stunting increased with the number of under-5 children in the household, with children in households of three or more facing a 40% increase in risk (OR: 1.40, 95% CI: 1.12–1.74). Conversely, maternal education and wealth acted as significant protective factors. Compared to those with no education, children of mothers with higher education had 59% lower odds of stunting (OR: 0.41, 95% CI: 0.30–0.57). Similarly, belonging to the richest wealth quintile significantly reduced the odds of stunting (OR: 0.57, 95% CI: 0.42–0.79).

Interestingly, the sex of the household head exhibited divergent effects on the two nutritional outcomes. Children in female-headed households had significantly lower odds of stunting (OR: 0.73, 95% CI: 0.59–0.89) but significantly higher odds of wasting (OR: 1.40, 95% CI: 1.03–1.92) compared to male-headed households. This suggests that while female-headed households may be more effective at managing long-term nutritional stability, they may be more vulnerable to acute shocks. Notably, after adjusting for spatial effects and other covariates, place of residence (urban/rural) and breastfeeding status did not reach statistical significance, as their credible intervals crossed the null value of one.

### Nonlinear Effects of Child and Maternal Age

The posterior mean estimates and 95% credible intervals for the nonlinear effects of child and maternal age are depicted in Figure 2. The model reveals a distinct, non-monotonic relationship between child age and nutritional status. For stunting (a), the risk increases sharply during the first 24 months of life, reflecting the critical first 1,000 days window where cumulative growth deficits typically manifest. After approximately 24 months, the effect plateaus, suggesting a stabilization of chronic growth failure as the child transitions to older age groups. In contrast, the risk of wasting shows a slight increase in early infancy but remains relatively more stable across the age spectrum compared to stunting, though the widening credible intervals at higher ages reflect increased posterior uncertainty.

**Figure 2.**
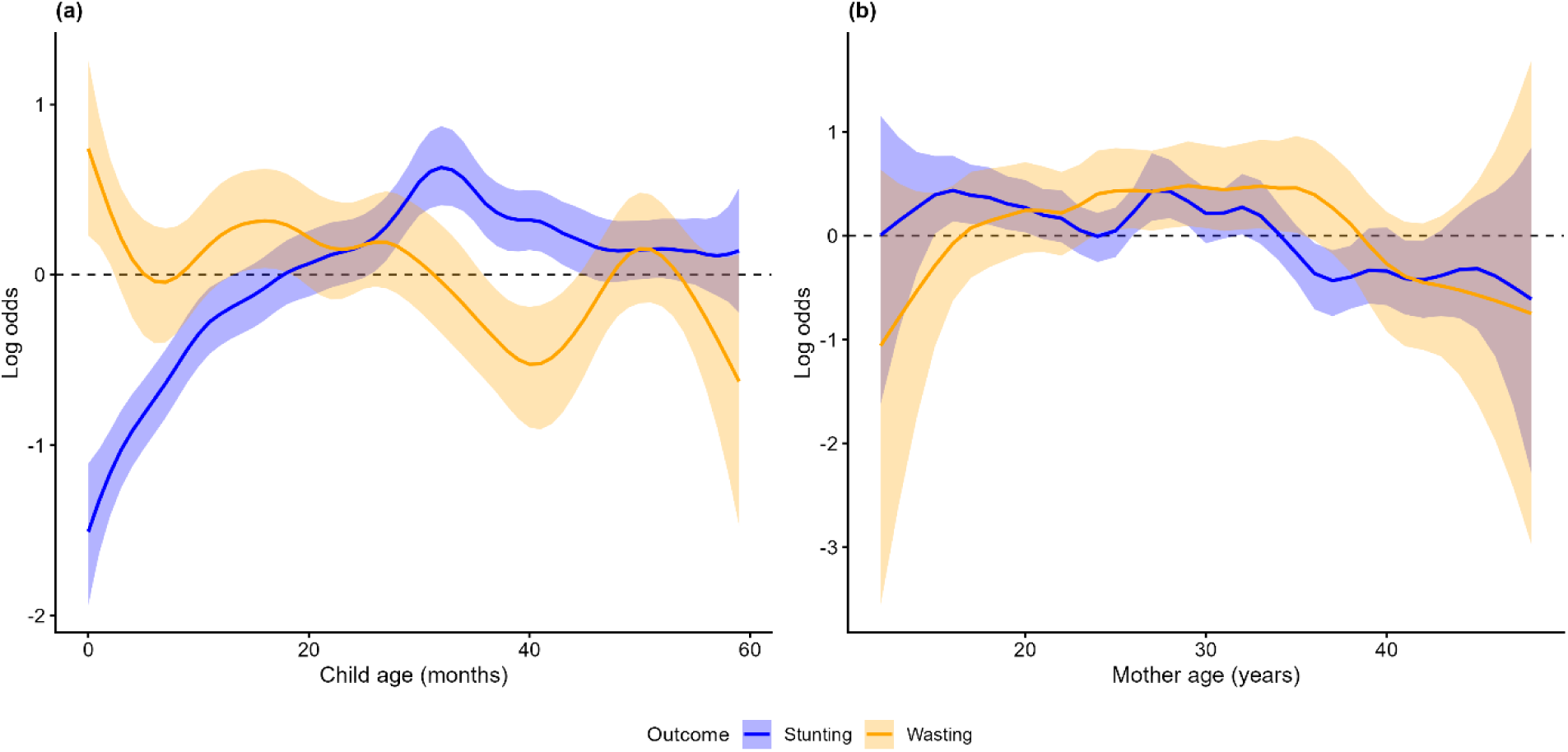
Outcome-specific effects of child and maternal age on stunting and wasting. (a) Child age in months and (b) maternal age in years. Solid lines and shaded areas represent posterior means and 95% credible intervals, respectively.

Regarding maternal age (b), the model identifies a nonlinear “U-shaped” or curvilinear trend, particularly for wasting. The risk of malnutrition is notably higher among children of very young mothers (adolescent mothers) and decreases as maternal age increases towards the late 20s and early 30s, likely due to increased maternal experience and improved household resource stability. However, there is a marginal increase in risk observed at the upper end of the maternal age distribution. By utilizing a random walk order 2 (RW2) prior for these continuous variables, the bivariate spatial model successfully accounts for these smooth, unconstrained functional forms, providing a more nuanced understanding of age-related vulnerabilities than traditional categorical or linear approaches.

The posterior mean estimates of the outcome-specific spatial random effects, derived from the bivariate SPDE model, are illustrated in Figure 3. These maps visualize the residual spatial variation, effectively isolating the unobserved geographic risk factors for stunting and wasting after adjusting for the child, maternal, and socio-economic predictors detailed in the fixed-effects analysis. In these visualizations, areas highlighted in red signify a higher-than-average residual risk locations where the observed malnutrition prevalence exceeds what is predicted by the model’s covariates. For stunting (a), the model identifies a prominent area of high residual risk concentrated across the northern and central highlands. Conversely, the residual risk for wasting (b) exhibits a distinct geographic signature, with higher-than-average risk identified in the northeastern pastoralist regions and specific localized areas in the south.

**Figure 3.**
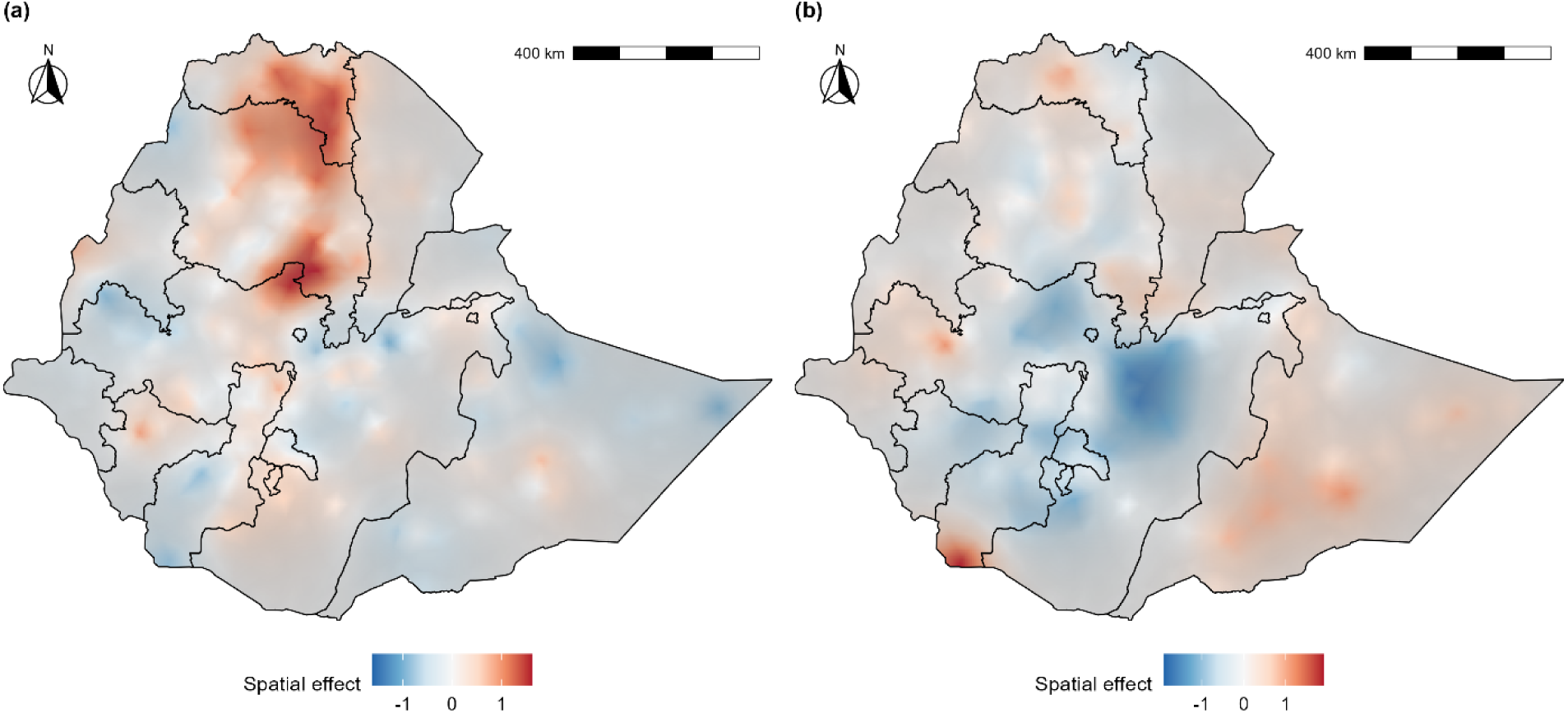
Posterior mean of the outcome-specific spatial random effects from the bivariate SPDE model for (a) stunting and (b) wasting among children under five in Ethiopia. The maps represent residual spatial variation after adjusting for covariates. Red areas indicate higher-than-average residual risk, while blue areas indicate lower-than-average risk. Semi-transparent gray shading denotes posterior uncertainty, with darker areas reflecting higher uncertainty, typically in regions with sparse cluster coverage.

Blue regions represent areas where the observed malnutrition is lower than predicted by the fixed effects, suggesting the presence of localized protective factors or regional health interventions not captured by the study variables. To ensure statistical rigor, semi-transparent gray shading is utilised to denote posterior uncertainty, with darker areas reflecting higher uncertainty typically found in regions with sparse cluster coverage in the EDHS 2019 dataset. This explicit mapping of uncertainty, combined with the identified nominal spatial range of 158 km, provides a more nuanced understanding of the underlying spatial processes than the raw exploratory maps. By decomposing the risk in this manner, the model highlights that even after neutralizing the effects of wealth and education, significant geographic disparities remain that necessitate targeted regional public health strategies.

### Posterior Mean Prevalence Estimates

The cluster-level posterior mean prevalence estimates, derived from the full bivariate spatial model (Model 4), are depicted in Figure 4. By synthesizing information from socio-economic covariates, non-linear age trends, and residual spatial dependency, the model provides a smoothed geographic risk profile that effectively mitigates the sampling noise inherent in raw, cluster-level data. Regarding the geographic variation in stunting (Figure 4a), the model identifies a high-prevalence corridor for chronic malnutrition extending across the northern and central highlands. In these regions, the smoothed posterior mean estimates for stunting consistently exceed the 40% critical threshold, particularly in clusters within the Amhara and Tigray regions.

**Figure 4.**
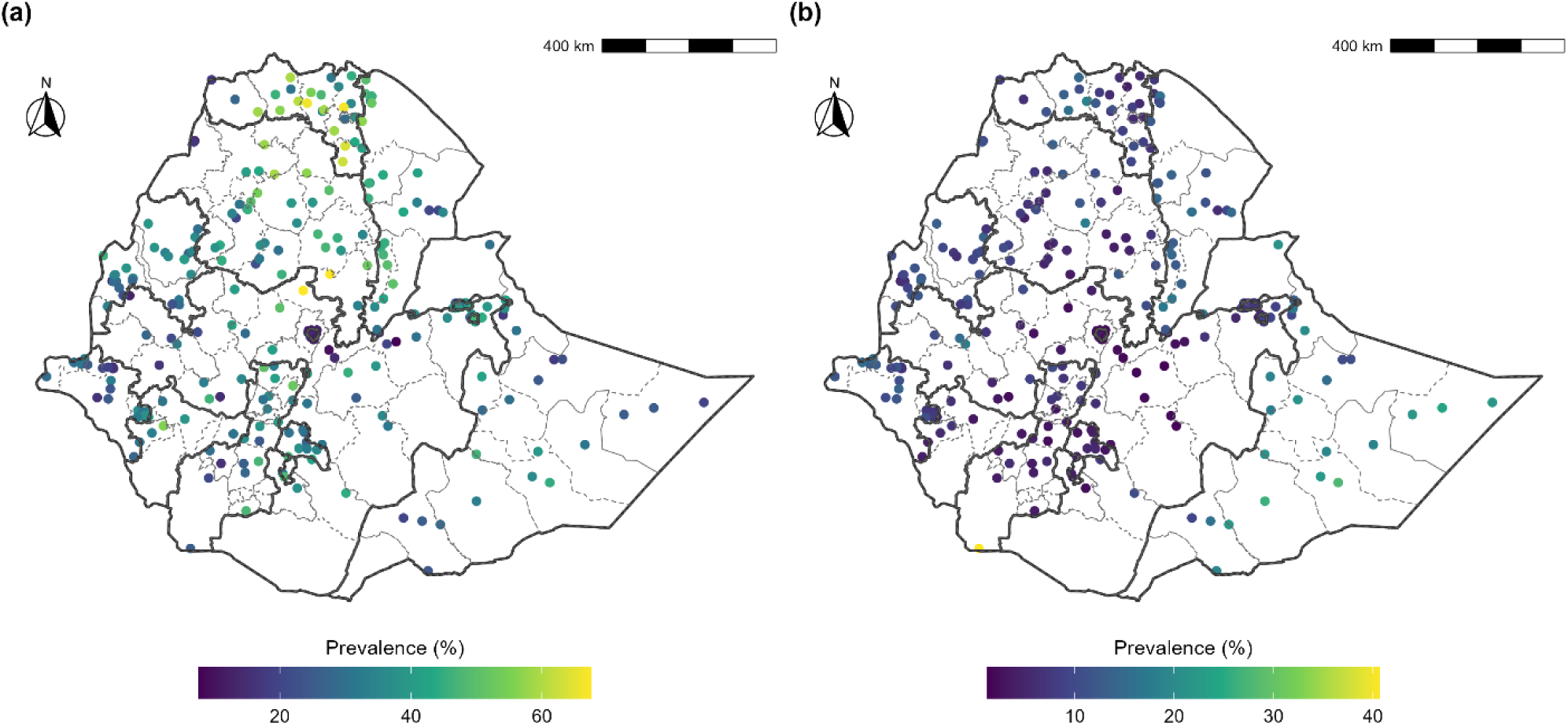
Spatial patterns of cluster-level posterior mean prevalence estimates from the Bayesian model in Ethiopia: (a) stunting and (b) wasting.

The risk of acute malnutrition follows a distinct spatial signature, as shown in the predicted prevalence for wasting (Figure 4b). The highest risk is concentrated in the northeastern pastoralist areas of the Afar region and parts of the Somali region, where posterior mean estimates frequently surpass the 15% threshold. The predictive clarity of these maps demonstrates the framework’s ability to handle geographic sparsity; by utilizing the estimated spatial range of 158 km, the SPDE approach produces stable and biologically plausible risk estimates even in areas with fewer survey clusters. Collectively, these results emphasize that while individual and household-level factors are significant drivers, the substantial geographic disparities remaining in the final smoothed maps necessitate region-specific public health interventions tailored to the distinct burdens of chronic and acute malnutrition.

### Exceedance Probabilities and Risk Assessment

To provide a formal statistical assessment of malnutrition risk relative to international benchmarks, Figure 5 presents the cluster-level exceedance probabilities derived from the posterior distribution. This metric quantifies the localized probability that malnutrition prevalence surpasses specific WHO-defined public health thresholds, offering a more robust measure of risk than point estimates alone. Regarding chronic malnutrition (Figure 5a), the map illustrates a nearly certain risk (probability approaching 1) that stunting prevalence exceeds the 20% threshold across the vast majority of clusters in the northern, central, and southwestern regions.

**Figure 5.**
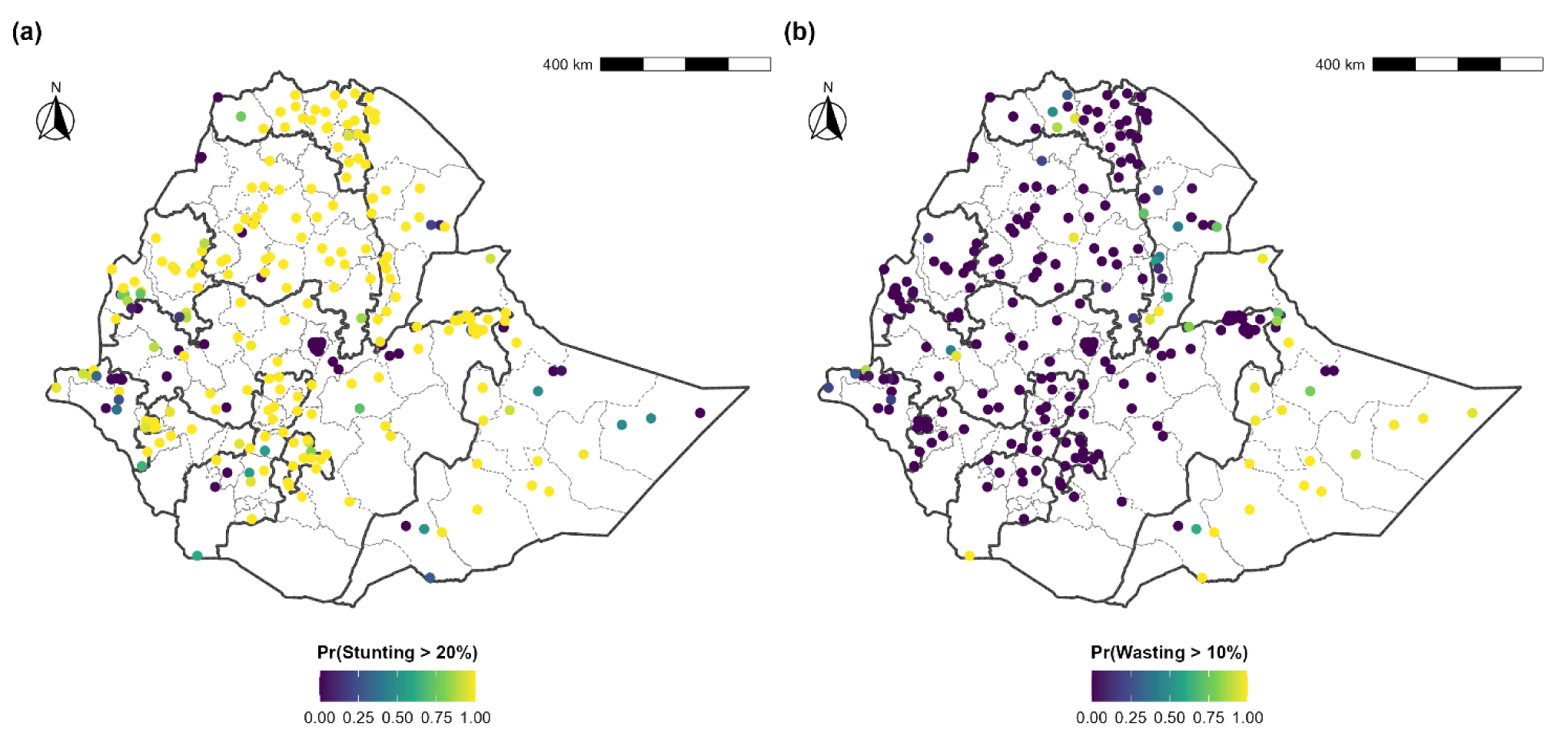
Spatial patterns of cluster-level exceedance probabilities in Ethiopia: (a) probability that stunting prevalence exceeds 20% and (b) probability that wasting prevalence exceeds 10%.

A more geographically concentrated risk profile is evident for acute malnutrition (Figure 5b), which displays the probability that wasting prevalence exceeds the 10% threshold. High exceedance probabilities are primarily clustered in the northeastern pastoralist zones of the Afar region and eastern segments of the Somali region, where many clusters show a probability greater than 0.75 of surpassing the threshold. In contrast, the probability remains low across much of the central and western regions. These exceedance maps serve as a critical decision-support tool, allowing policymakers to identify high-priority areas where there is a high statistical certainty that the nutritional burden has reached critical public health levels.

### Zonal Hotspot Analysis and Policy Implications

To facilitate targeted public health interventions, the cluster-level posterior estimates were aggregated to the administrative zonal level and classified according to WHO prevalence severity thresholds (Figure 6). The zonal-level analysis reinforces the presence of a substantial nutritional crisis, particularly regarding chronic malnutrition. For stunting (Figure 6a), the majority of administrative zones in the northern and central highlands, along with parts of the southwest, fall into the “Very High” severity category (>30%). These “hotspot” zones represent a contiguous belt of chronic growth failure that requires long-term structural investments in food security and maternal health.

**Figure 6.**
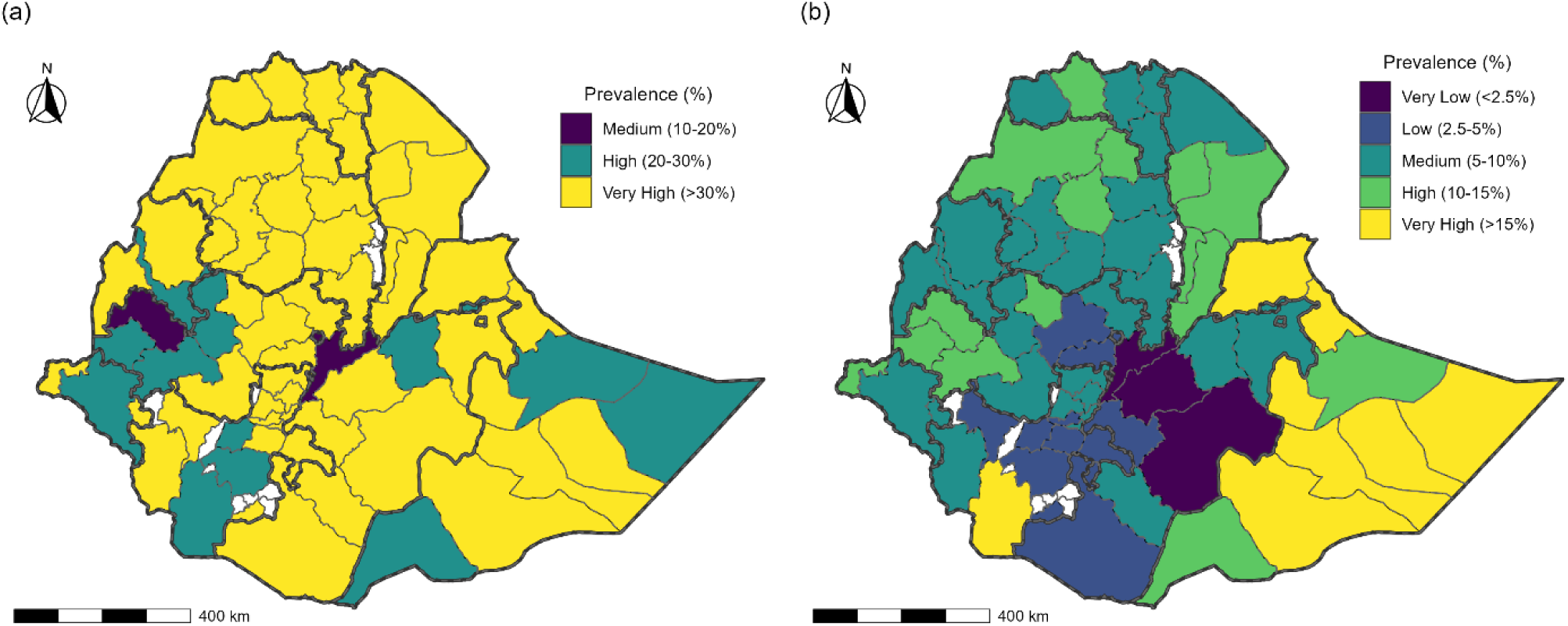
Hotspot zones of stunting and wasting in Ethiopia based on WHO prevalence classification

The distribution of wasting hotspots (Figure 6b) follows a more distinct pastoralist-driven pattern. “Very High” severity zones (>15%) are primarily concentrated in the northeastern Afar region and eastern Somali region. Furthermore, several “High” severity zones (10% - 15%) were identified across the northern and southern fringes of the country. Notably, while some central regions exhibit lower acute malnutrition rates, the widespread “Medium” (5% - 10%) and “High” classifications across the map underscore that wasting remains a persistent challenge beyond the primary hotspots. By identifying these specific administrative zones with high statistical certainty, this bivariate spatial framework provides a rigorous evidentiary basis for the prioritization of resources in Ethiopia’s national nutrition programmes.

## Discussion

This study provides robust empirical evidence that chronic (stunting) and acute (wasting) child malnutrition in Ethiopia are related yet spatially and etiologically distinct phenomena (3,4). By applying a Bayesian bivariate spatial (SPDE-INLA) framework to nationally representative EMDHS 2019 data, we move beyond conventional siloed analyses and demonstrate how joint modelling uncovers both shared and outcome-specific drivers that remain hidden in univariate approaches. The findings underscore the importance of decoupling long-term structural deprivation from short-term nutritional shocks when designing effective child nutrition policies.

Although stunting and wasting are biologically interconnected, the weak and statistically non-significant residual spatial correlation observed in this study suggests that, after accounting for shared socio-economic determinants, the two conditions follow largely independent geographic risk processes (6,9). This result aligns with emerging evidence that while wasting episodes can contribute to growth faltering, the translation of acute insults into chronic outcomes is heavily mediated by contextual factors such as food security, caregiving practices, and access to health services. The absence of strong residual spatial coupling implies that interventions targeting wasting hotspots may not automatically reduce stunting unless long-term structural determinants are simultaneously addressed.

The bivariate framework also highlights the analytical advantage of joint modelling. By allowing information sharing across outcomes, the model yields more stable estimates for wasting an inherently volatile condition while preserving outcome-specific associations. This methodological contribution is particularly relevant for low- and middle-income settings, where survey sparsity and geographic heterogeneity often limit inference for acute malnutrition indicators.

Consistent with previous studies in Ethiopia and sub-Saharan Africa, male children in our study were more likely than females to experience both stunting and wasting (7,8). This higher vulnerability may reflect a combination of biological susceptibility and differences in caregiving practices between boys and girls. Furthermore, children from multiple births faced a substantially increased risk, likely due to slower growth in the womb and greater competition for nutrition and care after birth.”

Maternal education and household wealth emerged as strong protective factors, particularly for stunting, reinforcing evidence that chronic malnutrition reflects cumulative socioeconomic deprivation (2,5). These findings reinforce the conceptualization of chronic malnutrition as a manifestation of cumulative socio-economic disadvantage. In contrast, the weaker and less consistent associations observed for wasting suggest that acute malnutrition is more sensitive to transient shocks such as illness, seasonal food scarcity, or climatic stress than to long-term household characteristics alone.

A notable and policy-relevant finding is the divergent role of female-headed households, echoing findings that women’s control over resources improves long-term child welfare while acute vulnerability persists under economic stress (20,25). While children in these households were less likely to be stunted, they faced higher odds of wasting. This pattern suggests that female-headed households may be effective in sustaining long-term nutritional investments but remain disproportionately exposed to acute economic or environmental shocks. Such households may benefit from targeted safety-net programmes designed to buffer short-term food insecurity.

The nonlinear effects of child age clearly delineate the first 24 months of life as the most critical period for the development of stunting, consistent with the established “first 1,000 days” framework (19,20). The sharp rise in risk during this window corroborates the well-established “first 1,000 days” paradigm and emphasizes the irreversibility of chronic growth failure once this period has passed. In contrast, the relatively flat age profile for wasting highlights its episodic nature and suggests opportunities for recovery if timely interventions are implemented.

The observed U-shaped relationship between maternal age and stunting risk further emphasizes the vulnerability of children born to adolescent mothers, likely reflecting biological immaturity, limited autonomy, and reduced access to resources. The slight increase in risk at older maternal ages may be attributable to parity-related resource dilution or declining maternal health, underscoring the need for age-sensitive maternal nutrition strategies.

Even after adjusting for a comprehensive set of covariates, substantial residual spatial heterogeneity persisted for both outcomes. The estimated spatial range of approximately 158 km indicates that unobserved drivers of malnutrition operate at a broad regional scale, potentially reflecting agro-ecological conditions, market access, or regional health system performance.

The spatial decoupling observed in the posterior risk maps has direct policy relevance and aligns with prior spatial analyses demonstrating distinct geographic processes for chronic versus acute undernutrition in Ethiopia (6,10). Chronic malnutrition hotspots concentrated in the northern and central highlands call for sustained investments in maternal education, agricultural diversification, and poverty alleviation. In contrast, wasting hotspots in the Afar and Somali regions areas characterised by pastoralist livelihoods and climatic volatility require rapid-response nutrition programmes, drought-resilient food systems, and mobile health services tailored to nomadic populations.

The exceedance probability maps further strengthen the policy utility of this framework by quantifying statistical certainty around WHO severity thresholds. This probabilistic approach offers a more defensible basis for prioritizing zones under Ethiopia’s national nutrition programmes than reliance on raw prevalence estimates alone.

The strength of this study lies in its integrated methodological approach, combining bivariate outcome modelling with advanced spatial inference under a Bayesian framework, consistent with best practice in Bayesian disease mapping and spatial epidemiology (12,13,24). The use of SPDE-INLA enables high-resolution risk mapping while appropriately accounting for uncertainty and spatial dependence. Incorporation of survey weights further enhances the representativeness of the findings.

Nevertheless, several limitations merit consideration, many of which are common to spatial analyses of DHS data (15,16). The cross-sectional design precludes causal inference and limits the ability to capture temporal dynamics, particularly seasonal variation in wasting. Additionally, DHS cluster displacement for confidentiality may introduce spatial misalignment, potentially attenuating fine-scale spatial patterns. Finally, unmeasured factors such as dietary diversity, micronutrient intake, and local health service quality could not be explicitly modelled and may contribute to residual spatial variation.

## Conclusion

This study demonstrates that while stunting and wasting share common socio-economic roots, their geographic expressions and immediate drivers differ substantially across Ethiopia. Treating these conditions as interchangeable outcomes risks misdirecting scarce public health resources. A dual strategy is therefore required: long-term, structurally oriented interventions to address chronic malnutrition in highland regions, alongside agile, shock-responsive programmes to mitigate acute malnutrition in pastoralist and climatically vulnerable areas.

By decoupling chronic and acute child malnutrition through a bivariate spatial lens, this work provides a rigorous evidence base to support geographically targeted, outcome-specific nutrition policies. Such precision is essential if Ethiopia is to accelerate progress towards its national nutrition targets and the Sustainable Development Goals.

## Acknowledgement

The author gratefully acknowledges the Demographic and Health Surveys (DHS) Programme for providing access to the data used in this study. The DHS Programme is internationally recognized for collecting and disseminating high-quality, nationally representative data on fertility, family planning, maternal and child health, gender, HIV/AIDS, malaria, and nutrition. The views expressed in this article are those of the author and do not necessarily reflect the views of the DHS Programme, the Ethiopia Demographic and Health Survey (EDHS), or the funding agencies.

## Author contributions

H.Y. Conceptualization, Methodology, Formal Analysis, Data Curation, Writing Original Draft Preparation, Review & Editing.

## Data availability

The EDHS data are publicly available and can be accessed from https://www.dhsprogramme.com/

## Declarations

### Ethics approval

Not applicable.

### Consent for publication

Not applicable.

### Clinical trial number

Not applicable

## Reference

1. UNICEF, WHO, World Bank Group. Levels and Trends in Child Malnutrition: Key Findings of the 2025 Edition [Internet]. 2025. Available from: https://data.unicef.org/resources/jme/

2. Woldekidan MA, Arja A, Worku G, Walker A, Kassebaum NJ, Hailemariam A, et al. The burden and trends of child and maternal malnutrition across the regions in Ethiopia, 1990–2019: The Global Burden of Disease Study 2019. PLOS Glob Public Heal [Internet]. 2024;4(7):1–17. Available from: 10.1371/journal.pgph.0002640

3. Sahiledengle B, Agho KE, Petrucka P, Kumie A, Beressa G, Atlaw D, et al. Concurrent wasting and stunting among under-five children in the context of Ethiopia: A generalised mixed-effects modelling. Matern \& Child Nutr. 2023;19(2).

4. Akombi BJ, Agho KE, Hall JJ, Wali N, Renzaho AMN, Merom D. Stunting, wasting and underweight in sub-Saharan Africa: a systematic review. Int J Environ Res Public Health. 2017;14(8):863.

5. Takele K, Zewotir T, Ndanguza D. Understanding correlates of child stunting in Ethiopia using generalized linear mixed models. BMC Public Health. 2019;19:626.

6. Kuse KA, Debeko DD. Spatial distribution and determinants of stunting, wasting and underweight in children under-five in Ethiopia. BMC Public Health. 2023;23:641.

7. Hiruy AF, Xiong Q, Jin Q, Zhao J, Lin X, He S, et al. The Association of Feeding Practices and Sociodemographic Factors on Underweight and Wasting in Children in Ethiopia: A Secondary Analysis of Four Health Surveys from 2000 to 2016. J Trop Pediatr [Internet]. 2021 Aug 27;67(4). Available from: https://academic.oup.com/tropej/article/doi/10.1093/tropej/fmab047/6358692

8. Gebru FK, others. Determinants of stunting among under-five children in Ethiopia: a multilevel mixed-effects analysis of 2016 Ethiopian demographic and health survey data. BMC Pediatr. 2019;19:176.

9. Kassie GW, Workie DL. Exploring the association of anthropometric indicators for under-five children in Ethiopia. BMC Public Health. 2019;19:764.

10. Dibisa KE, others. Prevalence and factors associated with acute malnutrition among children aged 6--59 months in West Wollega, Oromia, Ethiopia, 2024. J Heal Popul Nutr. 2025;44(1):319.

11. Louzada F, Nascimento DC do, Egbon OA. Spatial statistical models: An overview under the Bayesian approach. Axioms. 2021;10:307.

12. Best N, Richardson S, Thomson A. A comparison of Bayesian spatial models for disease mapping. Stat Methods Med Res. 2005;14(1):35–59.

13. Krainski E, Gómez-Rubio V, Bakka H, Lenzi A, Castro-Camilo D, Simpson D, et al. Advanced Spatial Modeling with Stochastic Partial Differential Equations Using {R} and {INLA}. Boca Raton, FL: Chapman and Hall/CRC; 2018.

14. Okango E, Mwambi H, Ngesa O. Spatial modeling of HIV and HSV-2 among women in Kenya with spatially varying coefficients. BMC Public Health. 2016;16(1):355.

15. Wakefield J. Disease mapping and spatial regression with count data. Biostatistics. 2007;8(2):158–83.

16. Lawson AB. Bayesian Disease Mapping: Hierarchical Modeling in Spatial Epidemiology. Boca Raton, FL: Chapman and Hall/CRC; 2018.

17. Ethiopia Public Health Institute (EPHI), ICF. Ethiopia Mini Demographic and Health Survey 2019: Final Report. Rockville, MD; 2021.

18. ICF International. Demographic and Health Survey Sampling and Household Listing Manual. Calverton, MD, USA; 2012.

19. World Health Organization. WHO Child Growth Standards: Length/Height-for-Age, Weight-for-Age, Weight-for-Length, Weight-for-Height and Body Mass Index-for-Age: Methods and Development. Geneva, Switzerland: World Health Organization; 2006.

20. UNICEF. UNICEF Conceptual Framework on Maternal and Child Nutrition. 2022.

21. Marshall EC, Spiegelhalter DJ. Identifying outliers in Bayesian hierarchical models: a simulation-based approach. In: Bayesian Statistics 8. Oxford University Press; 2007. p. 409–44.

22. Lang S, Brezger A. Bayesian P-splines. J Comput Graph Stat. 2004;13(1):183–212.

23. Goldstein H. Multilevel Statistical Models. Chichester, UK: John Wiley \& Sons; 2011.

24. Lindgren F, Rue H, Lindström J. An explicit link between Gaussian fields and Gaussian Markov random fields: the stochastic partial differential equation approach. J R Stat Soc Ser B (Statistical Methodol. 2011;73(4):423–98.

25. Pfeffermann D. The role of sampling weights when modeling survey data. Int Stat Rev. 1993;317–37.

26. Riebler A, Sørbye SH, Simpson D, Rue H. An intuitive Bayesian spatial model for disease mapping that accounts for scaling. Stat Methods Med Res. 2016;25(4):1145–65.

27. Chopin N. Approximate Bayesian inference for latent Gaussian models by using integrated nested Laplace approximations. J R Stat Soc Ser B (Statistical Methodol. 2009;71:319–92.

28. Simpson D, Rue H, Riebler A, Martins TG, Sørbye SH. Penalising model component complexity: A principled, practical approach to constructing priors. Stat Sci. 2017;32(1):1–28.

